# Growing up in poverty, growing old with multimorbidity in America, Britain, China, Europe and Indonesia: a retrospective and prospective study in 31 countries

**DOI:** 10.1101/2024.07.16.24310497

**Authors:** Gindo Tampubolon

**Affiliations:** NIHR Policy Research Unit on Healthy Ageing

**Keywords:** childhood poverty, multimorbidity, life course, fixed effects, America, Britain, China, Europe, Indonesia

## Abstract

The childhood poor in rich countries have reported worse cognitive, muscle and mental functions in old age. But it is uncertain whether the childhood poor around the world are at higher risks of multimorbidity because childhood recollections are often erroneous.

**METHODS:** Some 81,804 older adults over 50 years in 31 countries recalled their childhood conditions at ten to fourteen and prospectively reported their chronic conditions. Childhood conditions in Britain and Europe include numbers of books, rooms and people, presence of running hot water and central heating. Across in America, these are mostly replaced with financial hardship or family indebtedness, in China starvation to death due to government edict while in Indonesia presence of running cold water. Per prior practice childhood poverty is a latent construct of these error-laced recollections while multimorbidity is defined as at least two from a set of chronic conditions. Associations with multimorbidity are obtained with fixed effects probit model, controlling for age, education, wealth, marital status and youth illness. Extensive sensitivity analyses assessed robustness.

**RESULTS:** Childhood poverty associates with higher probabilities of multimorbidity (0.088, [95% confidence interval 0.057 – 0.118]) in 31 rich and developing countries. And women reported lower probabilities of multimorbidity (−0.071, [-0.096 – -0.047]). Frequencies of multimorbidity vary across countries, and likewise age profiles of multimorbidity in later life.

**DISCUSSION:** Evidence is accumulating that childhood lasts a life time in rich and developing nations. The various childhood recollections indicating poverty show that the childhood poor grow old with more chronic conditions. Because population ageing is posing a momentous challenge across the world, more research is needed on the life course shaping of healthy ageing. But this strong evidence calls for urgent actions to eliminate child poverty on account of its lifelong rewards. [287 + 4233 words]

## Introduction

Childhood poverty is a risk factor for old age disability, dysfunction and disease in America, Britain, China and Europe.(1–8) It raises question whether childhood poverty also marks multimorbidity – two or more chronic conditions – which often entails adverse clinical outcomes, high costs of care and mortality, not least due to unoptimized treatment that is geared toward single-condition treatment. (9,10) Ongoing ageing surveys have provided ample materials to examine the reach of childhood poverty.(11,12) Twenty eight trans-Atlantic countries from America through Britain to Israel report life course links that persist, marking cognitive and muscle function and depression status of older adults over 50 years who grew up as children in poverty.(3) With such comparative design admitting many countries, new evidence on life course shaping of multimorbidity can be used to advance efforts during the UN Decade of Healthy Ageing.(13) This is urgent because multimorbidity reduces older adults’ wellbeing and simultaneously increases costs of treatment and care placed on the patients, families and societies.(9,14,15)

This work aims to test the *hypothesis of life course shaping of multimorbidity* in America, Britain, China, Europe and Indonesia which posits that childhood poverty continues to mark multimorbidity in old age in 31 rich and developing nations even after adolescent health is considered – the lifelong association between poverty and multimorbidity remains. To achieve this five sister studies are used in a cross-country fixed effect design following earlier studies.(3,4) These are the Health & Retirement Study – HRS, English Longitudinal Study of Ageing – ELSA, Survey of Health Ageing & Retirement in Europe – SHARE, China Ageing, Health and Retirement Longitudinal Study – CHARLS, Indonesia Family Life Survey – IFLS.(16–20)

These 31 nations are chosen to encompass the span of ageing experience and its size: while Britain took eight decades to double the proportion of its 60 plus population, China only needs two decades to face the same predicament, a rapid velocity. Moreover these nations epitomise the momentous challenge posed by the ageing population: today this older population in China is fifteen times the size of its peer in Britain. But by the end of this decade according to the UN, it will be nineteen times at 370 million (https://population.un.org/wpp/ accessed 21 June 2024). With such velocity and mass, the momentum of ageing is challenging the world.

To understand older adults’ health, the frame of social determinants of health with links across the life course is adopted, defined as “conditions in the environments in which people are born and *raised*, live, learn, work, play, worship, and *age* that affect a wide range of health, functioning, and quality-of-life outcomes”.(21–23) The life course shaping hypothesis posits how one was *raised* materially has influence on the chance of experiencing multimorbidity as one *ages*.

When testing the hypothesis different concepts and indicators of childhood conditions are adopted in practice. Childhood poverty, childhood socioeconomic position and adverse childhood experience were some of them. Their indicators were often captured as scales or latent constructs. These indicators vary including parental social class, education and employment, self-report of family financial hardship or bankruptcy; being raised in a foster family; material facilities of childhood home such as number of rooms and people (indicating overcrowding), availability of books, running hot and cold water, central heating or indoor lavatory.(12,24–31) This study does not use the adverse childhood experience scale for two reasons. First, the scale is complementary to the other concepts. Second, the scale has more psychological or subjective elements which suggests more susceptibility to errors. I am not aware of any test of the magnitude of the errors, for instance by comparing a report of emotional abuse obtained during childhood with another report of the same experience obtained in later life, unlike childhood poverty which has undergone such veracity test in Britain.(1,2) Another veracity test of the childhood conditions recollection in a developing nation will be presented shortly. In sum, in rich and developing nations childhood recollection is erroneous, thus following prior practice a latent construct of childhood poverty is used.(1–5,7)

By examining the life course shaping hypothesis in rich and developing nations, large and small, we support efforts to secure healthy ageing including in the <u>UN Decade of Healthy Ageing,</u> through evidence on the risk factor of childhood poverty.(13)

## Methods and materials

### Outcome: multimorbidity

Following extensive empirical literature, a person has multimorbidity if two or more chronic conditions from a standard set are present; none and one chronic condition are considered together as the reference i.e. no multimorbidity; altogether making multimorbidity a binary outcome. The set lists diabetes, high cholesterol or dyslipidaemia, high blood pressure or hypertension, heart failure, stroke, chronic obstructive pulmonary disease, cancer of any except skin kind, and memory problems. This set was present in all five sister surveys.

### Childhood information

The participants were asked retrospectively about their childhood conditions up to eight decades earlier, then prospectively about their health. Retrospective indicators of childhood poverty, obtained from adults with average age of 66 year, raise concern about recall error. For example, in 2008 some 2500 Britons aged 50 were asked to recall the numbers of rooms and people in their homes when they were eleven (to assess overcrowding). Only one in three got both numbers right. Their mothers gave the objective answers when visited 39 years earlier; the families are Britain’s National Child Development Study 1958 cohort.(1,2) Similarly, in 2015 all adult members of the IFLS (wave 5) were asked the same two questions and water sources (cold running water, rain water etc.) when they were twelve. Of course, in 1993 (wave 1) the members were visited in their homes so matching can be made in order to check the answers – there were 1877 matches (aged twelve in 1993 and 34 in 2015). Fewer than one in ten (6%) got both numbers right. The best recalled is water sources (62%). In both rich and developing countries adults aged 34 – 50 years gave more erroneous answers than correct ones. It is scarcely plausible that an average 66 year old participant achieved perfect recall of childhood conditions. Assuming no errors in these retrospective surveys is unsafe.

Recent studies have responded to this concern by devising latent construct for childhood poverty, a practice followed here.(1–7) The alternative of using retrospective indicators as if they are perfectly recalled leads not only to unsafe inferences, it also prevents cross-country learning. For example, both Si and colleagues and Li and colleagues used childhood conditions collected in CHARLS to explain health in older Chinese.(8,32) The inclusion of death from starvation during the Great Leap Forward complicates comparison with other countries’ experience because no other countries in the HRS, ELSA, SHARE or IFLS, imposed comparable edict. Similarly, the presence of running *hot* water indicator in ELSA and SHARE, *were it included as is*, will equally complicate the comparison because this was not asked in CHARLS. To overcome this researchers from America, Britain and Europe have devised latent construct for childhood poverty which enables it to be used as a risk factor to explain multimorbidity in older adults. (1–7)

ELSA collected life history in wave 3, while SHARE collected childhood information in its life history waves (3 and 7 for those who did not provide it the first time). The HRS, ELSA and SHARE indicators have been described elsewhere.(2,3,33,34) HRS collected childhood information in the core interview at each wave but, crucially, this is not closely comparable with indicators from the other side of the Atlantic, instead opting to elaborate on financial situation. This information is augmented with the ad hoc Life History Mail Survey 2017. Such indicators were also used in the precursor of SHARE, the Survey of Income and Living Conditions in Europe which has been analysed with the same method used here.(12) CHARLS and IFLS both collected life history information in 2014. Although ageing studies in America, Britain, China, Europe and Indonesia have close family resemblance, the differences are worth listing (Table 1). (2,3,33,34)

**Table 1.** Retrospective childhood information in HRS, ELSA, CHARLS, SHARE & IFLS.

| Surveys | HRS (age <16) | ELSA & SHARE (age 10) |
| --- | --- | --- |
| Variable | Number of rooms<br>Number of people<br>Number of books<br>Ever experienced:<br>being financially poor had to move or<br>be helped because of financial<br>difficulty, had to live with grandparents,<br>father unemployed | Number of rooms<br>Number of people<br>Number of books<br>Physical facilities:<br>indoor toilet, fixed bath,<br>central heating, hot & cold<br>running water |
| Surveys | CHARLS (age <17) | IFLS (age 12) |
| Variable | Family member starved to death<br>Had to move due of financial difficulty<br>Experienced financial hardship<br>Living in commune that is unsafe<br>... less willing to help others<br>... not close knit<br>... not clean | Number of rooms<br>Number of people<br>Had electricity<br>Had cold running water<br>Had indoor toilet<br>Experienced hunger or famine |

### Non-response patterns for retrospective interviews

The analytic sample comprises prospective sample members who agreed to retrospective interviews. Some declined and there may be significant differences between the analytic *vs* declined sample, tested using x_2_ and *t* tests for nominal and continuous variables as appropriate (table 2). In the HRS the analytic sample has 61% females and 39% males, with x_2_ = 15.7 and p value < 0.001; a pattern also observed in IFLS. In contrast, in SHARE, ELSA and CHARLS analytic samples females are no more likely to agree to retrospective interviews. Meanwhile in the HRS the analytic sample has older members and this difference is significant while in SHARE, ELSA and IFLS the opposite is true while also significant; CHARLS is found in between, with no significant difference. In sum, there are no strong patterns of participation in retrospective interviews around the world – a notable finding.

**Table 2.**
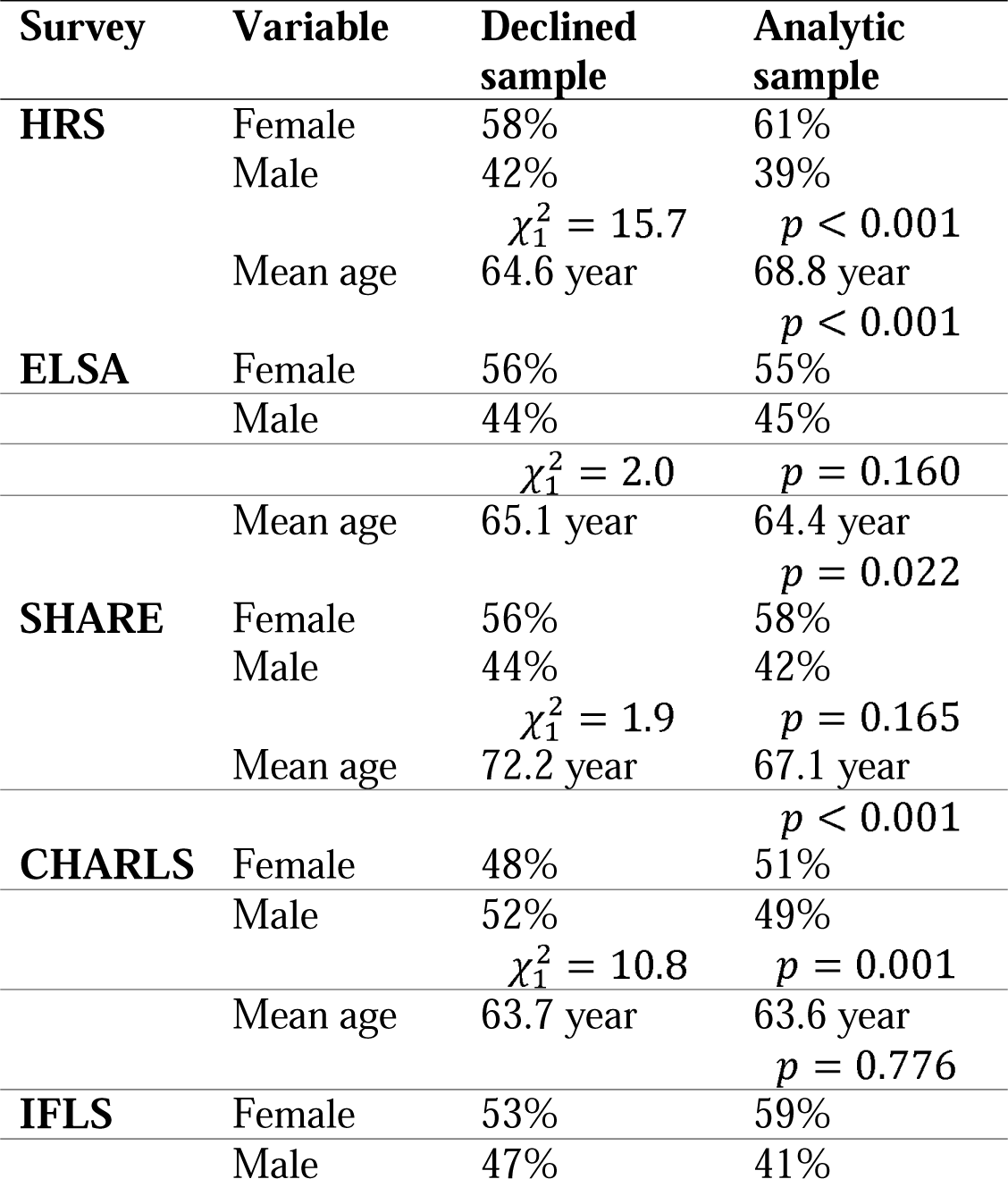
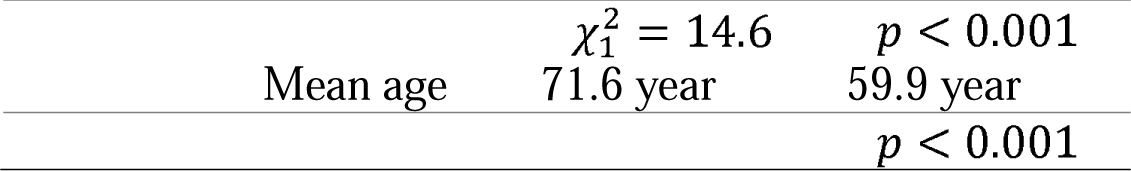
Non-response patterns across analytic *vs* declined sample in HRS, ELSA, CHARLS, SHARE & IFLS.

| Survey | Variable | Declined sample | Analytic sample |
| --- | --- | --- | --- |
| **HRS** | Female | 58% | 61% |
|  | Male | 42% | 39% |
| | | $\chi^2_1 = 15.7$ | $p < 0.001$ |
| | Mean age | 64.6 year | 68.8 year<br>$p < 0.001$ |
| **ELSA** | Female | 56% | 55% |
|  | Male | 44% | 45% |
| | | $\chi^2_1 = 2.0$ | $p = 0.160$ |
| | Mean age | 65.1 year | 64.4 year<br>$p = 0.022$ |
| **SHARE** | Female | 56% | 58% |
|  | Male | 44% | 42% |
| | | $\chi^2_1 = 1.9$ | $p = 0.165$ |
| | Mean age | 72.2 year | 67.1 year<br>$p < 0.001$ |
| **CHARLS** | Female | 48% | 51% |
|  | Male | 52% | 49% |
| | | $\chi^2_1 = 10.8$ | $p = 0.001$ |
| | Mean age | 63.7 year | 63.6 year<br>$p = 0.776$ |
| **IFLS** | Female | 53% | 59% |
|  | Male | 47% | 41% |

|  |  |  |
| --- | --- | --- |
| | $\chi^2_1 = 14.6$ | $p < 0.001$ |
| Mean age | 71.6 year | 59.9 year |
| | | $p < 0.001$ |

To test the life course shaping hypothesis, the latent variable of childhood poverty was matched with multimorbidity in 2020/2021 except for CHARLS and IFLS in 2014/2015. A set of social determinants is included as standard in the literature including age, sex, youth illness, education, marital status and wealth (2,3,5,35,36) Per prior practice studying America, Britain and Europe, all analyses control for differences between countries by including country fixed effects, and standard error correction was applied because the key exposure of childhood poverty is an estimated latent variable.(3,37) To check robustness of the results *sensitivity analyses* were conducted: random effects instead of fixed effects, gender-stratified analysis, alternative threshold of multimorbidity (three or more), count of chronic conditions with Poisson model (See Supplement). Statistical significance was set at 5% (95% confidence interval/CI) and Latent GOLD version 6 was used for estimation.(38)

## Results

Table 3 collects features of the analytic sample which has 56% female and mean age 65.7 year. Among female participants 19.7% reported more multimorbidity; among males this figure is 20%. Importantly, one in five experienced poverty in childhood, giving power to estimation in fully adjusted model.

**Table 3.**
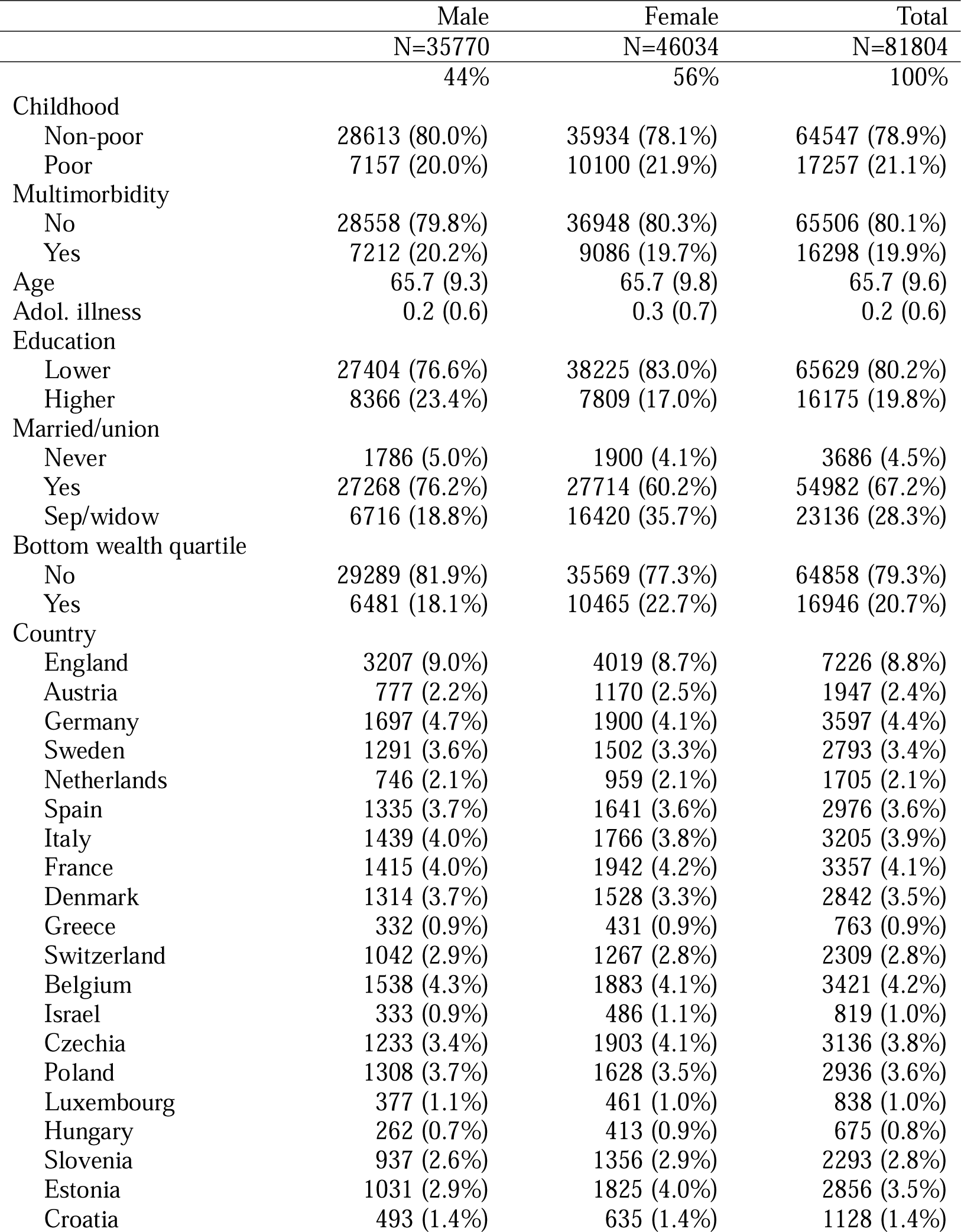
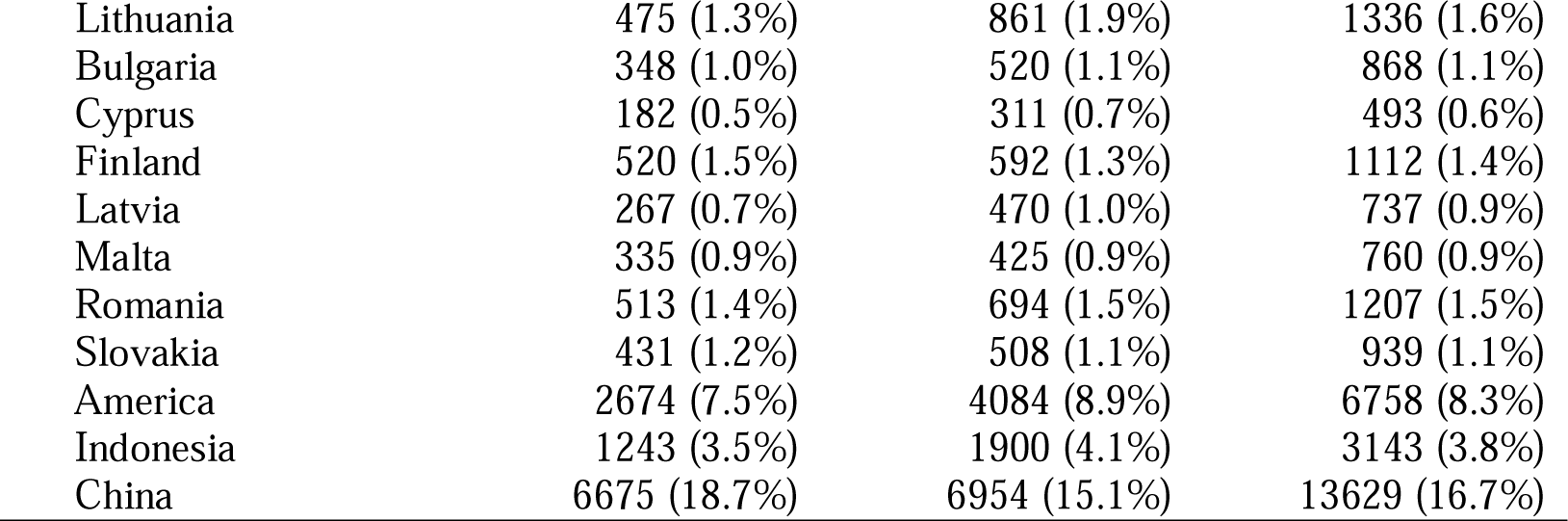
Summary features of the analytic sample with categorical variables in numbers (percentages) and continuous variables in means (standard deviations). Sources HRS, ELSA, CHARLS, SHARE and IFLS.

|  | Male | Female | Total |
| --- | --- | --- | --- |
|  | N=35770 | N=46034 | N=81804 |
|  | 44% | 56% | 100% |
| Childhood |  |  |  |
| Non-poor | 28613 (80.0%) | 35934 (78.1%) | 64547 (78.9%) |
| Poor | 7157 (20.0%) | 10100 (21.9%) | 17257 (21.1%) |
| Multimorbidity |  |  |  |
| No | 28558 (79.8%) | 36948 (80.3%) | 65506 (80.1%) |
| Yes | 7212 (20.2%) | 9086 (19.7%) | 16298 (19.9%) |
| Age | 65.7 (9.3) | 65.7 (9.8) | 65.7 (9.6) |
| Adol. illness | 0.2 (0.6) | 0.3 (0.7) | 0.2 (0.6) |
| Education |  |  |  |
| Lower | 27404 (76.6%) | 38225 (83.0%) | 65629 (80.2%) |
| Higher | 8366 (23.4%) | 7809 (17.0%) | 16175 (19.8%) |
| Married/union |  |  |  |
| Never | 1786 (5.0%) | 1900 (4.1%) | 3686 (4.5%) |
| Yes | 27268 (76.2%) | 27714 (60.2%) | 54982 (67.2%) |
| Sep/widow | 6716 (18.8%) | 16420 (35.7%) | 23136 (28.3%) |
| Bottom wealth quartile |  |  |  |
| No | 29289 (81.9%) | 35569 (77.3%) | 64858 (79.3%) |
| Yes | 6481 (18.1%) | 10465 (22.7%) | 16946 (20.7%) |
| Country |  |  |  |
| England | 3207 (9.0%) | 4019 (8.7%) | 7226 (8.8%) |
| Austria | 777 (2.2%) | 1170 (2.5%) | 1947 (2.4%) |
| Germany | 1697 (4.7%) | 1900 (4.1%) | 3597 (4.4%) |
| Sweden | 1291 (3.6%) | 1502 (3.3%) | 2793 (3.4%) |
| Netherlands | 746 (2.1%) | 959 (2.1%) | 1705 (2.1%) |
| Spain | 1335 (3.7%) | 1641 (3.6%) | 2976 (3.6%) |
| Italy | 1439 (4.0%) | 1766 (3.8%) | 3205 (3.9%) |
| France | 1415 (4.0%) | 1942 (4.2%) | 3357 (4.1%) |
| Denmark | 1314 (3.7%) | 1528 (3.3%) | 2842 (3.5%) |
| Greece | 332 (0.9%) | 431 (0.9%) | 763 (0.9%) |
| Switzerland | 1042 (2.9%) | 1267 (2.8%) | 2309 (2.8%) |
| Belgium | 1538 (4.3%) | 1883 (4.1%) | 3421 (4.2%) |
| Israel | 333 (0.9%) | 486 (1.1%) | 819 (1.0%) |
| Czechia | 1233 (3.4%) | 1903 (4.1%) | 3136 (3.8%) |
| Poland | 1308 (3.7%) | 1628 (3.5%) | 2936 (3.6%) |
| Luxembourg | 377 (1.1%) | 461 (1.0%) | 838 (1.0%) |
| Hungary | 262 (0.7%) | 413 (0.9%) | 675 (0.8%) |
| Slovenia | 937 (2.6%) | 1356 (2.9%) | 2293 (2.8%) |
| Estonia | 1031 (2.9%) | 1825 (4.0%) | 2856 (3.5%) |
| Croatia | 493 (1.4%) | 635 (1.4%) | 1128 (1.4%) |
| Lithuania | 475 (1.3%) | 861 (1.9%) | 1336 (1.6%) |
| Bulgaria | 348 (1.0%) | 520 (1.1%) | 868 (1.1%) |
| Cyprus | 182 (0.5%) | 311 (0.7%) | 493 (0.6%) |
| Finland | 520 (1.5%) | 592 (1.3%) | 1112 (1.4%) |
| Latvia | 267 (0.7%) | 470 (1.0%) | 737 (0.9%) |
| Malta | 335 (0.9%) | 425 (0.9%) | 760 (0.9%) |
| Romania | 513 (1.4%) | 694 (1.5%) | 1207 (1.5%) |
| Slovakia | 431 (1.2%) | 508 (1.1%) | 939 (1.1%) |
| America | 2674 (7.5%) | 4084 (8.9%) | 6758 (8.3%) |
| Indonesia | 1243 (3.5%) | 1900 (4.1%) | 3143 (3.8%) |
| China | 6675 (18.7%) | 6954 (15.1%) | 13629 (16.7%) |

### Multimorbidity profile across countries

Most older adults reported none or one chronic condition, except in America. Of those with two or more chronic conditions they are distributed differently across the national sample, compare for instance Estonia, Latvia and China in the first column. Together these reflect the histories and health systems variations across these nations, from the NHS in Britain to the largely private health system in America. The implications will be discussed shortly, comprising China and America as illustration.

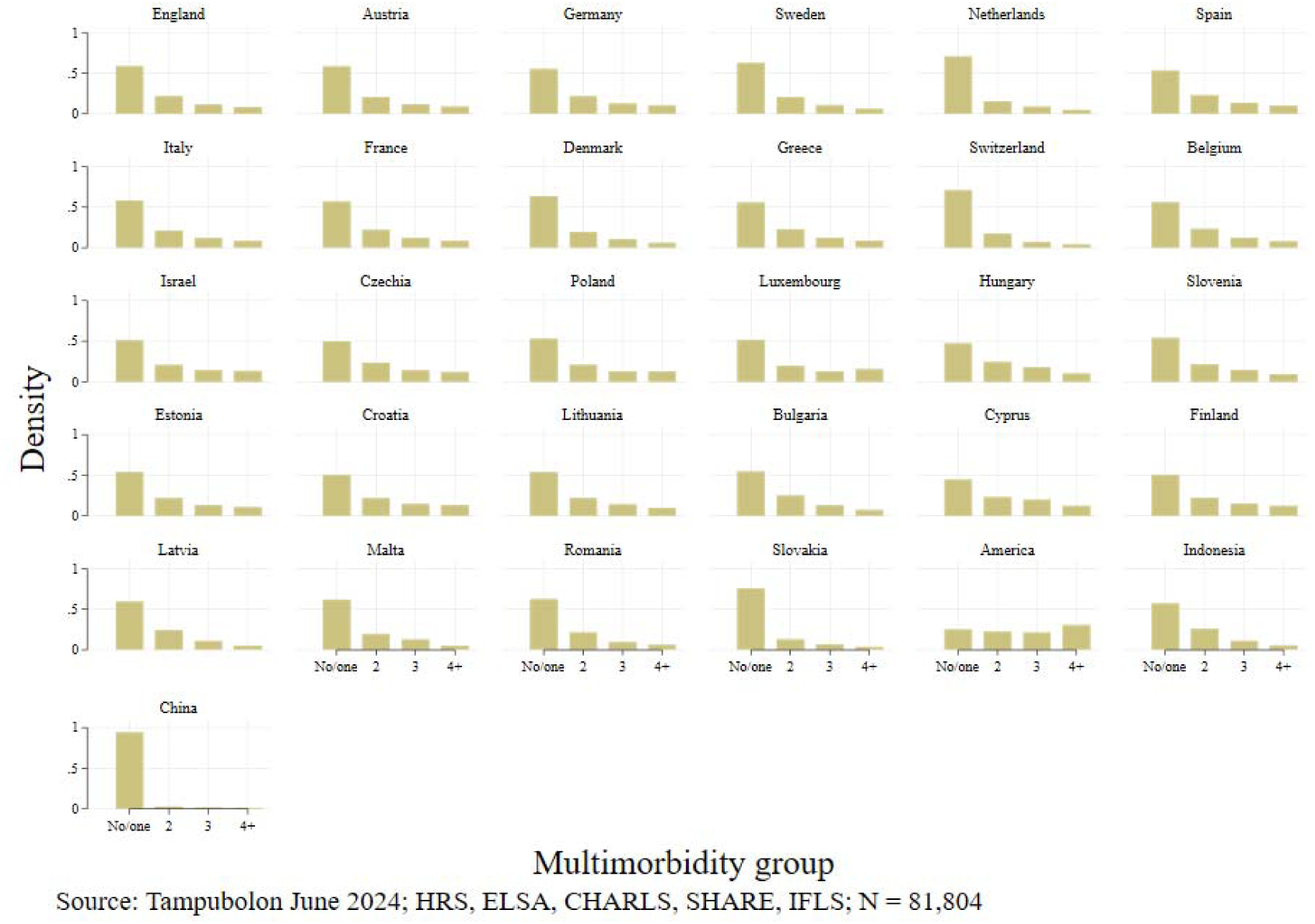

The probit model coefficients explaining multimorbidity are collected in table 4 which shows that in America, Britain, China, Europe and Indonesia growing up in poverty accompanies growing old with multimorbidity (coefficient 0.088, *p* ≤0.001), females report lower probability of multimorbidity (−0.071, *p*≤0.001) and the probabilities increase with age (0.027, *p*≤0.001). Briefly, the other covariates suggest that college education (compared to high school or lower) associates with lower probabilities of multimorbidity (−0.189, *p*≤0.001). Being in the bottom third of wealth distribution associates with higher risks of multimorbidity. Importantly, reflecting the social determinants framework above, illness during adolescence is significant and in the expected direction (0.229, *p*≤0.001). This leaves childhood poverty coefficient to be interpreted as a fully adjusted association with multimorbidity.

**Table 4.** Probit coefficients explaining multimorbidity in HRS, ELSA, CHARLS, SHARE and IFLS. Constant includes the reference country Britain.

| Variable | Coef. | Std.Err. | P | Lower CI | Upper CI |
| --- | --- | --- | --- | --- | --- |
| Constant | -6.876 | 0.356 | <0.001 | -7.573 | -6.179 |
| Childhood poor | 0.088 | 0.016 | <0.001 | 0.057 | 0.118 |
| Sex, F | -0.071 | 0.012 | <0.001 | -0.096 | -0.047 |
| Age | 0.027 | 0.001 | <0.001 | 0.026 | 0.029 |
| Higher educated | -0.189 | 0.015 | <0.001 | -0.219 | -0.159 |
| Never married | -0.046 | 0.030 | 0.130 | -0.106 | 0.013 |
| Married | 0.006 | 0.015 | 0.680 | -0.023 | 0.036 |
| Adol. illness | 0.229 | 0.009 | <0.001 | 0.211 | 0.247 |
| Bottom wealth quart. | 0.021 | 0.019 | 0.270 | -0.017 | 0.058 |
| Austria | 4.514 | 0.353 | <0.001 | 3.822 | 5.205 |
| Germany | 4.653 | 0.352 | <0.001 | 3.962 | 5.343 |
| Sweden | 4.347 | 0.352 | <0.001 | 3.657 | 5.038 |
| Netherlands | 4.287 | 0.353 | <0.001 | 3.595 | 4.979 |
| Spain | 4.463 | 0.352 | <0.001 | 3.773 | 5.153 |
| Italy | 4.480 | 0.352 | <0.001 | 3.790 | 5.171 |
| France | 4.464 | 0.352 | <0.001 | 3.774 | 5.154 |
| Denmark | 4.465 | 0.352 | <0.001 | 3.774 | 5.155 |
| Greece | 4.723 | 0.354 | <0.001 | 4.028 | 5.418 |
| Switzerland | 4.131 | 0.353 | <0.001 | 3.439 | 4.822 |
| Belgium | 4.551 | 0.352 | <0.001 | 3.861 | 5.241 |
| Israel | 4.764 | 0.354 | <0.001 | 4.069 | 5.458 |
| Czechia | 4.675 | 0.352 | <0.001 | 3.985 | 5.365 |
| Poland | 4.668 | 0.352 | <0.001 | 3.978 | 5.359 |
| Luxembourg | 4.705 | 0.354 | <0.001 | 4.011 | 5.400 |
| Hungary | 4.760 | 0.355 | <0.001 | 4.064 | 5.456 |
| Slovenia | 4.685 | 0.352 | <0.001 | 3.994 | 5.376 |
| Estonia | 4.560 | 0.352 | <0.001 | 3.870 | 5.251 |
| Croatia | 4.668 | 0.354 | <0.001 | 3.975 | 5.361 |
| Lithuania | 4.754 | 0.353 | <0.001 | 4.061 | 5.446 |
| Bulgaria | 4.631 | 0.354 | <0.001 | 3.936 | 5.325 |
| Cyprus | 4.775 | 0.356 | <0.001 | 4.077 | 5.472 |
| Finland | 4.773 | 0.354 | <0.001 | 4.080 | 5.466 |
| Latvia | 4.517 | 0.355 | <0.001 | 3.822 | 5.213 |
| Malta | 4.558 | 0.355 | <0.001 | 3.863 | 5.253 |
| Romania | 4.431 | 0.354 | <0.001 | 3.737 | 5.125 |
| Slovakia | 4.268 | 0.355 | <0.001 | 3.573 | 4.963 |
| America | -0.044 | 0.733 | 0.950 | -1.480 | 1.392 |
| Indonesia | 0.425 | 0.691 | 0.540 | -0.929 | 1.778 |
| China | 0.130 | 0.530 | 0.810 | -0.908 | 1.169 |

To focus on the childhood poverty, for each nation the predicted probabilities of multimorbidity over the age of 70 to 90 are plotted as distinguished by childhood poverty, then these plots are collected in a trellis plot (Figure 1). This shows at a glance how childhood poverty shapes multimorbidity in old age across a wide range of history and health systems in the rich and developing nations.

**Figure 1.**
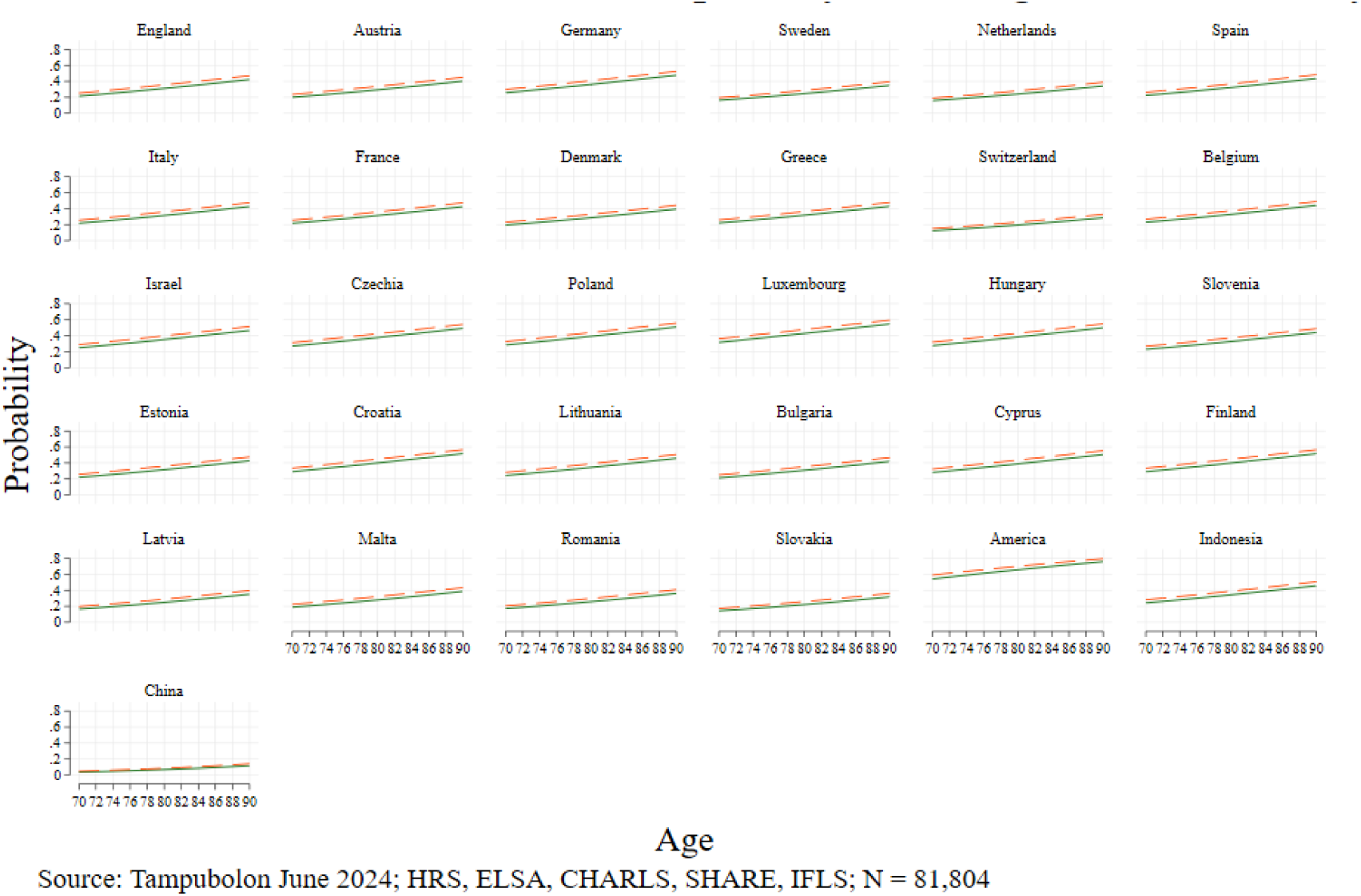
Probabilities of multimorbidity among the childhood poor (dash) and non-poor (solid) in older people aged 70 to 90 years in Britain, Europe and America based on models in table 3 where all covariates are set at the sample averages. Analysis of HRS, ELSA, CHARLS, SHARE and IFLS.

The trellis plot reveals several key patterns including ones obtained for the first time here. (Statistical significance has been presented above.) First, age is an important risk factor though it works differently in different countries, validating the fixed effect design. For example, compare the levels and slopes of Slovakia and America (penultimate row). Second, childhood poverty puts people at a disadvantage in old age, putting the dashed lines always above the solid lines. Third, the regional patterns of Europe are discernible especially distinguishing the Scandinavian and Eastern European nations. Together, the variations in levels and slopes are marked (e.g. the penultimate row without Indonesia), preventing one single summary pattern from representing all rich countries.

Finally, the contrast between America and China is instructive while counter intuitive. The comparison apparently suggests that there are more older Americans than Chinese with multimorbidity for the same age, more than half versus less than one in ten at 70. Moreover, the slope in America is steeper. America being richer than China in income terms should have suggested the opposite if the well-known wealth and health association founded our intuition. Instead the pattern is a manifestation of survival bias or selective mortality.(39) The samples in the two countries differ markedly, giving rise to this counter intuitive pattern, where in China the history of the Great Leap Forward left only the strongest to survive until very old ages of 70 to 90. This determines the lower level and gentler slope of multimorbidity in the country. This is perhaps coupled with the less sensitive or comprehensive diagnostics in the country compared to those in America with its largely private health system. Together these result in a marked counter intuitive pattern obtained here for the first time. Sensitivity analyses were conducted and the results above stand (Supplement).

**Table 5.**
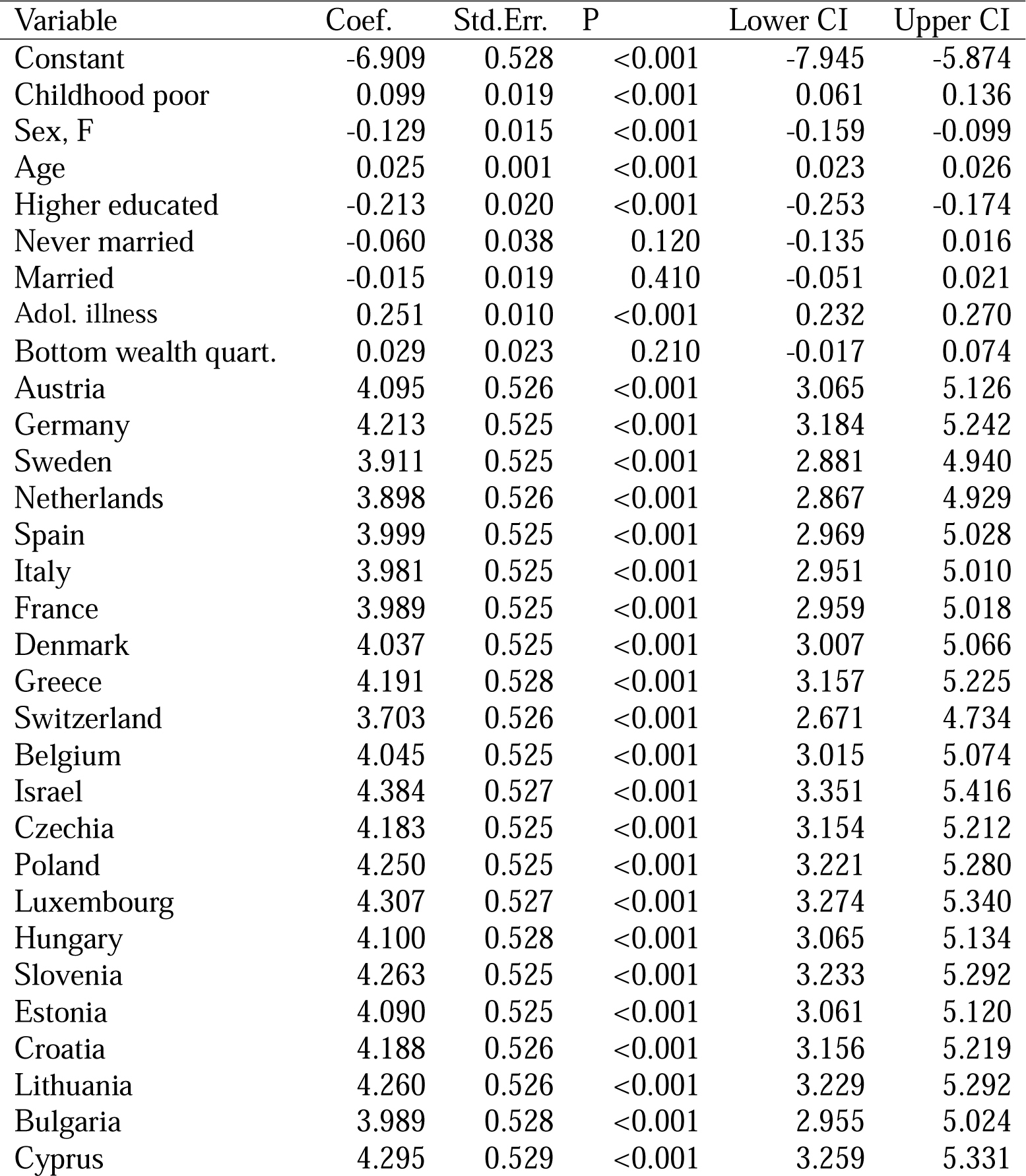
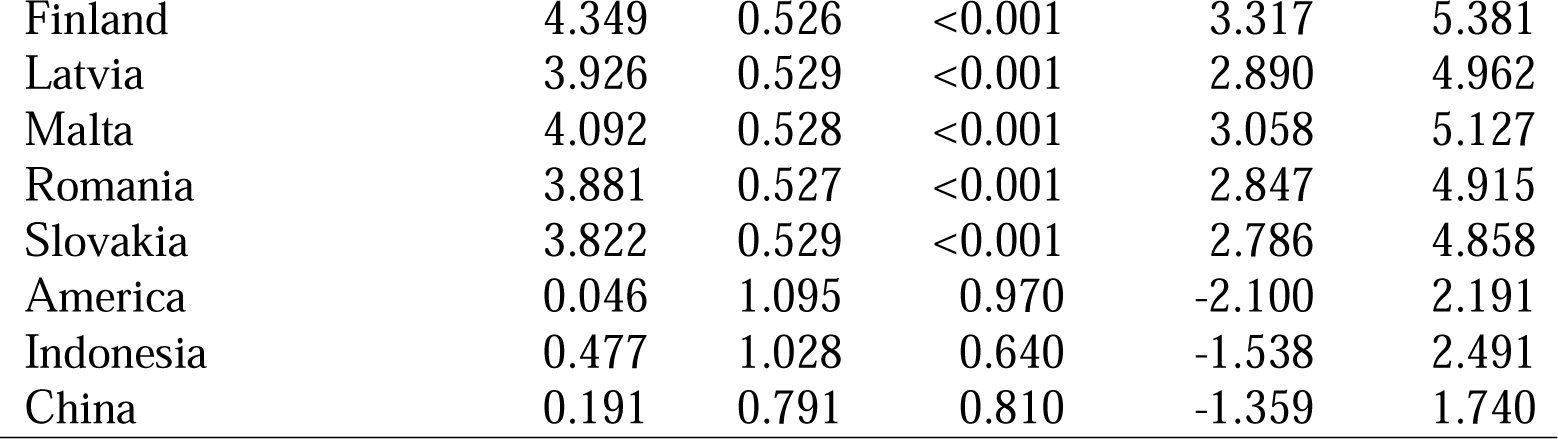
Sensitivity analysis: Probit coefficients with fixed effects explaining multimorbidity as three or more chronic conditions in America (HRS), Britain (ELSA), China (CHARLS), Europe (SHARE) and Indonesia (IFLS).

| Variable | Coef. | Std.Err. | P | Lower CI | Upper CI |
| --- | --- | --- | --- | --- | --- |
| Constant | -6.909 | 0.528 | <0.001 | -7.945 | -5.874 |
| Childhood poor | 0.099 | 0.019 | <0.001 | 0.061 | 0.136 |
| Sex, F | -0.129 | 0.015 | <0.001 | -0.159 | -0.099 |
| Age | 0.025 | 0.001 | <0.001 | 0.023 | 0.026 |
| Higher educated | -0.213 | 0.020 | <0.001 | -0.253 | -0.174 |
| Never married | -0.060 | 0.038 | 0.120 | -0.135 | 0.016 |
| Married | -0.015 | 0.019 | 0.410 | -0.051 | 0.021 |
| Adol. illness | 0.251 | 0.010 | <0.001 | 0.232 | 0.270 |
| Bottom wealth quart. | 0.029 | 0.023 | 0.210 | -0.017 | 0.074 |
| Austria | 4.095 | 0.526 | <0.001 | 3.065 | 5.126 |
| Germany | 4.213 | 0.525 | <0.001 | 3.184 | 5.242 |
| Sweden | 3.911 | 0.525 | <0.001 | 2.881 | 4.940 |
| Netherlands | 3.898 | 0.526 | <0.001 | 2.867 | 4.929 |
| Spain | 3.999 | 0.525 | <0.001 | 2.969 | 5.028 |
| Italy | 3.981 | 0.525 | <0.001 | 2.951 | 5.010 |
| France | 3.989 | 0.525 | <0.001 | 2.959 | 5.018 |
| Denmark | 4.037 | 0.525 | <0.001 | 3.007 | 5.066 |
| Greece | 4.191 | 0.528 | <0.001 | 3.157 | 5.225 |
| Switzerland | 3.703 | 0.526 | <0.001 | 2.671 | 4.734 |
| Belgium | 4.045 | 0.525 | <0.001 | 3.015 | 5.074 |
| Israel | 4.384 | 0.527 | <0.001 | 3.351 | 5.416 |
| Czechia | 4.183 | 0.525 | <0.001 | 3.154 | 5.212 |
| Poland | 4.250 | 0.525 | <0.001 | 3.221 | 5.280 |
| Luxembourg | 4.307 | 0.527 | <0.001 | 3.274 | 5.340 |
| Hungary | 4.100 | 0.528 | <0.001 | 3.065 | 5.134 |
| Slovenia | 4.263 | 0.525 | <0.001 | 3.233 | 5.292 |
| Estonia | 4.090 | 0.525 | <0.001 | 3.061 | 5.120 |
| Croatia | 4.188 | 0.526 | <0.001 | 3.156 | 5.219 |
| Lithuania | 4.260 | 0.526 | <0.001 | 3.229 | 5.292 |
| Bulgaria | 3.989 | 0.528 | <0.001 | 2.955 | 5.024 |
| Cyprus | 4.295 | 0.529 | <0.001 | 3.259 | 5.331 |
| Finland | 4.349 | 0.526 | <0.001 | 3.317 | 5.381 |
| Latvia | 3.926 | 0.529 | <0.001 | 2.890 | 4.962 |
| Malta | 4.092 | 0.528 | <0.001 | 3.058 | 5.127 |
| Romania | 3.881 | 0.527 | <0.001 | 2.847 | 4.915 |
| Slovakia | 3.822 | 0.529 | <0.001 | 2.786 | 4.858 |
| America | 0.046 | 1.095 | 0.970 | -2.100 | 2.191 |
| Indonesia | 0.477 | 1.028 | 0.640 | -1.538 | 2.491 |
| China | 0.191 | 0.791 | 0.810 | -1.359 | 1.740 |

**Table 6.** Poisson coefficients of multimorbidity (number of chronic conditions, fixed effects, 31 countries, N=81,804)

| Variable | Coef. | Std.Err. | P | Lower CI | Upper CI |
| --- | --- | --- | --- | --- | --- |
| Constant | -12.964 | 0.040 | <0.001 | -13.041 | -12.886 |
| Childhood poor | 0.066 | 0.011 | <0.001 | 0.045 | 0.088 |
| Sex, F | -0.075 | 0.009 | <0.001 | -0.092 | -0.058 |
| Age | 0.023 | 0.001 | <0.001 | 0.022 | 0.024 |
| Higher educated | -0.165 | 0.012 | <0.001 | -0.188 | -0.143 |
| Never married | -0.063 | 0.023 | 0.006 | -0.108 | -0.018 |
| Married | 0.005 | 0.011 | 0.650 | -0.016 | 0.026 |
| Adol. illness | 0.183 | 0.006 | <0.001 | 0.172 | 0.194 |
| Bottom wealth quart. | 0.014 | 0.013 | 0.270 | -0.011 | 0.040 |
| Austria | 11.501 | 0.030 | <0.001 | 11.442 | 11.560 |
| Germany | 11.611 | 0.026 | <0.001 | 11.561 | 11.661 |
| Sweden | 11.327 | 0.028 | <0.001 | 11.271 | 11.383 |
| Netherlands | 11.287 | 0.034 | <0.001 | 11.220 | 11.354 |
| Spain | 11.443 | 0.026 | <0.001 | 11.391 | 11.494 |
| Italy | 11.457 | 0.026 | <0.001 | 11.406 | 11.509 |
| France | 11.426 | 0.027 | <0.001 | 11.373 | 11.478 |
| Denmark | 11.389 | 0.029 | <0.001 | 11.331 | 11.446 |
| Greece | 11.622 | 0.036 | <0.001 | 11.552 | 11.693 |
| Switzerland | 11.111 | 0.033 | <0.001 | 11.047 | 11.176 |
| Belgium | 11.488 | 0.026 | <0.001 | 11.436 | 11.540 |
| Israel | 11.661 | 0.037 | <0.001 | 11.588 | 11.735 |
| Czechia | 11.608 | 0.025 | <0.001 | 11.558 | 11.658 |
| Poland | 11.611 | 0.027 | <0.001 | 11.557 | 11.664 |
| Luxembourg | 11.641 | 0.039 | <0.001 | 11.566 | 11.716 |
| Hungary | 11.639 | 0.037 | <0.001 | 11.567 | 11.712 |
| Slovenia | 11.615 | 0.027 | <0.001 | 11.562 | 11.668 |
| Estonia | 11.528 | 0.026 | <0.001 | 11.477 | 11.579 |
| Croatia | 11.595 | 0.033 | <0.001 | 11.530 | 11.660 |
| Lithuania | 11.661 | 0.032 | <0.001 | 11.599 | 11.724 |
| Bulgaria | 11.546 | 0.036 | <0.001 | 11.476 | 11.616 |
| Cyprus | 11.654 | 0.042 | <0.001 | 11.572 | 11.736 |
| Finland | 11.702 | 0.033 | <0.001 | 11.637 | 11.767 |
| Latvia | 11.472 | 0.040 | <0.001 | 11.394 | 11.549 |
| Malta | 11.537 | 0.038 | <0.001 | 11.462 | 11.612 |
| Romania | 11.428 | 0.036 | <0.001 | 11.357 | 11.498 |
| Slovakia | 11.124 | 0.049 | <0.001 | 11.028 | 11.219 |
| America | -1.408 | 0.029 | <0.001 | -1.465 | -1.351 |
| Indonesia | -0.107 | 0.036 | 0.003 | -0.177 | -0.037 |
| China | -0.973 | 0.025 | <0.001 | -1.023 | -0.924 |

**Table 7.** Probit coefficients of multimorbidity, Female (2 or more, fixed effects, 31 countries, N=81,804)

| Variable | Coef. | Std.Err. | P | Lower CI | Upper CI |
| --- | --- | --- | --- | --- | --- |
| Constant | -6.889 | 0.386 | <0.001 | -7.645 | -6.133 |
| Childhood poor | 0.112 | 0.021 | <0.001 | 0.071 | 0.153 |
| Age | 0.028 | 0.001 | <0.001 | 0.027 | 0.030 |
| Higher educated | -0.231 | 0.022 | <0.001 | -0.273 | -0.188 |
| Never married | -0.068 | 0.042 | 0.100 | -0.150 | 0.014 |
| Married | -0.006 | 0.019 | 0.740 | -0.043 | 0.030 |
| Adol. illness | 0.219 | 0.012 | <0.001 | 0.196 | 0.241 |
| Bottom wealth quart. | 0.057 | 0.024 | 0.018 | 0.010 | 0.105 |
| Austria | 4.402 | 0.381 | <0.001 | 3.655 | 5.148 |
| Germany | 4.486 | 0.380 | <0.001 | 3.741 | 5.231 |
| Sweden | 4.189 | 0.381 | <0.001 | 3.443 | 4.934 |
| Netherlands | 4.162 | 0.382 | <0.001 | 3.414 | 4.910 |
| Spain | 4.363 | 0.380 | <0.001 | 3.618 | 5.108 |
| Italy | 4.384 | 0.380 | <0.001 | 3.639 | 5.129 |
| France | 4.291 | 0.380 | <0.001 | 3.546 | 5.036 |
| Denmark | 4.354 | 0.381 | <0.001 | 3.608 | 5.100 |
| Greece | 4.548 | 0.384 | <0.001 | 3.795 | 5.300 |
| Switzerland | 3.905 | 0.381 | <0.001 | 3.158 | 4.653 |
| Belgium | 4.461 | 0.380 | <0.001 | 3.716 | 5.206 |
| Israel | 4.625 | 0.384 | <0.001 | 3.874 | 5.377 |
| Czechia | 4.546 | 0.380 | <0.001 | 3.801 | 5.290 |
| Poland | 4.606 | 0.381 | <0.001 | 3.860 | 5.352 |
| Luxembourg | 4.500 | 0.384 | <0.001 | 3.748 | 5.253 |
| Hungary | 4.673 | 0.384 | <0.001 | 3.919 | 5.426 |
| Slovenia | 4.521 | 0.381 | <0.001 | 3.775 | 5.266 |
| Estonia | 4.470 | 0.380 | <0.001 | 3.725 | 5.215 |
| Croatia | 4.604 | 0.382 | <0.001 | 3.855 | 5.353 |
| Lithuania | 4.721 | 0.382 | <0.001 | 3.973 | 5.469 |
| Bulgaria | 4.548 | 0.384 | <0.001 | 3.796 | 5.299 |
| Cyprus | 4.745 | 0.386 | <0.001 | 3.988 | 5.501 |
| Finland | 4.614 | 0.383 | <0.001 | 3.864 | 5.364 |
| Latvia | 4.465 | 0.384 | <0.001 | 3.712 | 5.217 |
| Malta | 4.514 | 0.384 | <0.001 | 3.761 | 5.267 |
| Romania | 4.348 | 0.383 | <0.001 | 3.598 | 5.099 |
| Slovakia | 4.223 | 0.384 | <0.001 | 3.470 | 4.976 |
| America | -0.070 | 0.780 | 0.930 | -1.600 | 1.459 |
| Indonesia | 0.453 | 0.730 | 0.530 | -0.977 | 1.883 |
| China | 0.112 | 0.594 | 0.850 | -1.052 | 1.275 |

**Table 8.** Probit coefficients of multimorbidity, Male (2 or more, fixed effects, 31 countries, N=81,804)

| Variable | Coef. | Std.Err. | P | Lower CI | Upper CI |
| --- | --- | --- | --- | --- | --- |
| Constant | -6.584 | 0.379 | <0.001 | -7.327 | -5.841 |
| Childhood poor | 0.054 | 0.024 | 0.025 | 0.007 | 0.100 |
| Age | 0.026 | 0.001 | <0.001 | 0.024 | 0.027 |
| Higher educated | -0.152 | 0.022 | <0.001 | -0.196 | -0.108 |
| Never married | 0.028 | 0.046 | 0.540 | -0.063 | 0.119 |
| Married | 0.070 | 0.027 | 0.010 | 0.017 | 0.122 |
| Adol. illness | 0.249 | 0.015 | <0.001 | 0.220 | 0.277 |
| Bottom wealth quart. | -0.069 | 0.032 | 0.030 | -0.131 | -0.007 |
| Austria | 4.292 | 0.373 | <0.001 | 3.561 | 5.022 |
| Germany | 4.488 | 0.371 | <0.001 | 3.760 | 5.215 |
| Sweden | 4.185 | 0.372 | <0.001 | 3.457 | 4.914 |
| Netherlands | 4.078 | 0.373 | <0.001 | 3.347 | 4.809 |
| Spain | 4.234 | 0.372 | <0.001 | 3.506 | 4.962 |
| Italy | 4.243 | 0.372 | <0.001 | 3.514 | 4.971 |
| France | 4.323 | 0.372 | <0.001 | 3.594 | 5.051 |
| Denmark | 4.247 | 0.372 | <0.001 | 3.518 | 4.976 |
| Greece | 4.581 | 0.376 | <0.001 | 3.844 | 5.319 |
| Switzerland | 4.030 | 0.373 | <0.001 | 3.300 | 4.761 |
| Belgium | 4.307 | 0.371 | <0.001 | 3.579 | 5.035 |
| Israel | 4.587 | 0.376 | <0.001 | 3.850 | 5.325 |
| Czechia | 4.482 | 0.372 | <0.001 | 3.754 | 5.211 |
| Poland | 4.408 | 0.372 | <0.001 | 3.679 | 5.138 |
| Luxembourg | 4.594 | 0.376 | <0.001 | 3.857 | 5.331 |
| Hungary | 4.502 | 0.379 | <0.001 | 3.761 | 5.244 |
| Slovenia | 4.533 | 0.372 | <0.001 | 3.804 | 5.262 |
| Estonia | 4.299 | 0.372 | <0.001 | 3.570 | 5.028 |
| Croatia | 4.397 | 0.375 | <0.001 | 3.663 | 5.131 |
| Lithuania | 4.401 | 0.375 | <0.001 | 3.666 | 5.135 |
| Bulgaria | 4.396 | 0.377 | <0.001 | 3.657 | 5.136 |
| Cyprus | 4.411 | 0.382 | <0.001 | 3.662 | 5.160 |
| Finland | 4.610 | 0.374 | <0.001 | 3.877 | 5.344 |
| Latvia | 4.186 | 0.380 | <0.001 | 3.441 | 4.930 |
| Malta | 4.248 | 0.377 | <0.001 | 3.509 | 4.987 |
| Romania | 4.199 | 0.376 | <0.001 | 3.462 | 4.935 |
| Slovakia | 3.964 | 0.377 | <0.001 | 3.225 | 4.703 |
| America | 0.001 | 0.769 | 0.990 | -1.506 | 1.508 |
| Indonesia | 0.372 | 0.733 | 0.610 | -1.064 | 1.808 |
| China | 0.136 | 0.531 | 0.800 | -0.904 | 1.177 |

**Table 9.** Probit coefficients of multimorbidity (2+, random effects, 31 countries, N=81,804)

| Variable | Coef. | Std.Err. | P | Lower CI | Upper CI |
| --- | --- | --- | --- | --- | --- |
| Constant | -2.682 | 0.081 | <0.001 | -2.841 | -2.524 |
| Childhood poor | 0.231 | 0.027 | <0.001 | 0.178 | 0.284 |
| Sex, F | -0.068 | 0.026 | 0.010 | -0.119 | -0.016 |
| Age | 0.028 | 0.001 | <0.001 | 0.026 | 0.030 |
| Higher educated | -0.195 | 0.023 | <0.001 | -0.240 | -0.151 |
| Never married | -0.047 | 0.034 | 0.160 | -0.114 | 0.019 |
| Married | 0.002 | 0.013 | 0.870 | -0.024 | 0.028 |
| Adol. illness | 0.229 | 0.011 | <0.001 | 0.206 | 0.251 |
| Bottom wealth quart. | 0.022 | 0.037 | 0.560 | -0.051 | 0.095 |

**Supplement Tables 6, 7, 8 and 9. Sensitivity analyses: Poisson coefficients explaining number of chronic conditions** (**6**)**; Probit coefficients with random effects** (**7**) **and stratified by sex (8 and 9), explaining multimorbidity in America (HRS), Britain (ELSA), China (CHARLS), Europe (SHARE) and Indonesia (IFLS)**

## Discussion: life course links and global links

Evidence is accumulating that the early life stage reaches into old age. Childhood poverty continues to shape the health of older people even in rich nations across many chronic conditions or multimorbidity. This is the first evidence from 31 nationwide representative surveys of how childhood poverty plays a role in multimorbidity variations in older adults over 50. This operates in diverse health systems: in a national health system (UK), a largely private health system (US) and everything in between. The British National Health Service has a history that spans more than 75 years old and has accompanied Britons in the study participants throughout their life courses. Using latent construct to recover child poverty, the analysis shows important associations between childhood poverty and multimorbidity that are consistently observed across the world, enriching our appreciation of the life course shaping of health in old age.

What mechanism lies behind this shaping is an important question. I have tried to offer an epigenetic explanation and evidence for the role of childhood poverty as a risk factor.(3) Poverty in childhood is found to induce epigenetic change through increasing methylation rates which leaves a mark that lasts until old age in the American Venous Blood Sample.(3,40,41) Children with similar genetic make-up may nevertheless differ in their phenotypic expression, hence organ function, because their childhood conditions differ in ways that accumulate adverse methylation rates i.e. epigenetic changes in one child more than in another.(3,42) That childhood lasts a lifetime, we knew. Now we know it also shapes the chance of multimorbidity.

What happens in one stage of life shapes health in much later stage, likewise what happens in one country shapes what can happen in other countries regarding older people with multimorbidity, more specifically their long term care. Recall China and Britain. If China requires social care workers for its older population at the end of this decade (nineteen times that of Britain), for long term care associated with multimorbidity, and China invited care workers from overseas, then both countries will compete for these workers. Today Britain is already struggling to attract social care workers from overseas, despite their own special visa lane.(43) The evidence here about the influence of childhood poverty on multimorbidity suggests that more older adults will need long term care, and the competition for social care workers is likely to get more intense.

Even with the preceding insights there are limits to what can be learned from this work. First, its framework is not a full structural framework,(4) although this has precedents.(2,5,44) That alternative has encouraged the authors to suggest that were more covariates considered, the association with childhood poverty would be entirely weakened. This suggestion is of course an eminently empirical question. The imperative remains that a fuller framework should be considered when all information on youth and adult conditions in all these countries are available. Second, this work is also limited in its focus on material lack in childhood, lacking psychological variables such as parent(adult)-child nurturing or abusive relationships. Cultivating psychological development is of course core to the life course stage of raising children.(45) Third, by including fixed effects this work controls for unobserved differences between nations such as their histories and health systems. Fourth, putting together these sister studies of HRS, SHARE, ELSA, CHARLS and IFLS which vary in their retrospective indicators may be a step too far. It is better to analyse each survey separately, one may suggest. Of course efforts to harmonise these sister studies (and more) are desirable and ongoing e.g. the Gateway to Aging initiative. This methodological approach of deriving a latent construct of childhood poverty complements such initiatives. Moreover, nation by nation analyses for these 31 nations can be done in future monographs. Last, a key limitation arises from the design being observational rather than a randomised study (to childhood material intervention and to control), preventing causal interpretation of these results. The childhood poor may differ from the non-poor in ways not observed. Moreover, some of the childhood poor may not have survived to take part in these longitudinal randomised ageing studies. Although this survivor effect may suggest a direction of bias, its magnitude is unknown. See the comparison between America and China above.

The strengths were mentioned throughout but three need emphasis. First, this is the first analysis to study 31 countries in the family of Health and Retirement Study, with closely comparable key exposure and health outcomes that speak directly to the UN Decade of Healthy Ageing.(13) The geographic reach of the sample as well as the variety of health systems it encompasses, together strengthen the generalisation of the results to nations around the world. Second, all the parallel surveys are ongoing, making this work a unique pad to launch future works on the consequences of child poverty in old age, on age trajectories of myriad health outcomes and multimorbidity in ageing populations across nations.

Another advantage of this frame of life course shaping which constructs childhood poverty as a latent variable is it can handle disparate information collected across ageing surveys as indicators of childhood poverty, from lack of running hot or cold water to financial hardship to starving to death due to government edict. In turn this means three things. For example: it is possible to encompass childhood war experience such as the Russian war in Ukraine and the Hamas-Israel war in Palestine. Second, it can uncover critical difference across the world such as survival effects in China. Had these 31 countries not been put into the same frame, we would not have noticed the critical consequence of world history. (3,46,47) Last, it offers parsimony without losing much theoretical insight along the way. Li et al 2020 use 11 childhood factors while Si et al add more, giving 13 factors, while giving similar results i.e. childhood disadvantage relates to worse health outcomes (more frailty and weaker intrinsic capacity, respectively). The framework can include unique factors such as starving to death due to government edict (Mao’s Great Leap Forward) which is historically specific because few countries have comparable experience since the mid-20^th^ century. This frame of life course shaping allows diverse and historically rooted childhood conditions to be explored as to their influences much later in life in many countries around the world, rich or otherwise.

### Reflection on recent literature

These findings echo recent studies in different countries though not all.(1–3,5–7,33,34) While consensus has not yet arrived, the evidence is largely supportive of a life course shaping of multimorbidity. The contrast with the new findings here can arise from the outcome studied or the special nature of the sample. Here nationally representative samples are used, comparable across 31 countries. Further, fixed effects capturing idiosyncratic country effects such as unique health system and history, are included. All agree in showing that the childhood poor experience more multimorbidity in old age.

But a few studies on the life course shaping of health in older ages have reported different results, which may be due to variations in variable construction, outcomes, methods, health systems and broader structures of societies. Vable and coauthors in America suggest that once socioeconomic mobility in adulthood is considered, no direct association remains between childhood economic status and old age health.(7) On this side of the Atlantic, studies in Sweden and Europe have also shown varying associations between poverty or economic status in childhood with old age health.(4–6) But the construct of childhood conditions vary. Lennartsson and colleagues depart from the usual practice of ignoring recall error by using latent construct of childhood poverty, in fact a full structural model to explain old age health indicated by pain, fatigue and breathing difficulty in Sweden; meanwhile Pakpahan and colleagues also used a structural model to explain self-rated health in old age in 13 European countries.(4,5)

In Sweden the authors suggest that childhood poverty is no longer binding on old age health once adult conditions are considered. This may be explained in two ways. The extensive welfare state of Sweden has been known to deliver exceptional health service to its older population. Our own work comparing 17 countries in Europe shows that Sweden has managed to break the link between economic position and sensory impairment.(48) Lennartsson and colleagues may have found a manifestation of Sweden’s exceptionalism in ameliorating pain, fatigue and breathing difficulty. But this can sit with the results above: on multimorbidity childhood poverty continues to matter.(5) Sweden’s exceptionalism may not have succeeded in breaking all the links between childhood poverty and the spectrum of health in old age. If indeed the mechanism involves epigenetic changes, amelioration is possible but perhaps the chance of complete elimination is slim.

It is worth demonstrating above that childhood recall is erroneous, putting empirical studies which ignore this onto unsafe inference.

### Future research and policy including the UN Decade of Healthy Ageing

Evidence from around the world laid here raises several implications for research and global health policy. First, retrospective childhood information is viable and can be fruitfully examined by recognising that this information is laced with error. Crucially, childhood poverty, a latent construct built on several retrospective indicators, can still be obtained.(1–3)

Second, this strong result from many countries speaks directly to global health policy such as the <u>UN Decade of Healthy Ageing</u>.(13) The initiative can grab the opportunity to emphasise the life course shaping of health in older ages around the world, showing the magnitudes and significance as obtained here. If rich countries with their advanced health systems have experienced this life course shaping of health, the many low and middle income countries are also likely to carry the long reach of childhood poverty. The 31 countries in this study are mostly high income countries; those who grew rich before growing older. China and Indonesia are the only middle income countries in the study.

In Britain the Chief Medical Officer recently reported on critical plans to ensure healthy ageing in the country.(10) The report entitled “Health in an Ageing Society” emphasises within-country spatial disparity in multimorbidity, in particular showing older adult with greater need of care are found in coastal and rural areas. This constitutes specific care and research challenges. Overseas, the low and middle-income countries are in a decidedly different predicament of growing old before growing rich. Moreover, the WHO observed that the countries with the difficult predicament are growing old faster.(49) These countries should be the focus of future research on the life course shaping of health in older ages.

The findings are uniquely relevant to healthy ageing policies to reduce the burden of multimorbidity in older age through keener appreciations of its deep roots, lending support to global initiatives that aim to support families and communities in raising children outside poverty. While more knowledge is needed, especially on how economic development entails epigenetic changes operating across the life course, strong evidence is at hand to help secure a decade of healthy ageing for the world.

## Ethical review

The University of Manchester exempted the investigation from full ethical review as it uses publicly available deidentified secondary datasets.

## STROBE list

STROBE list is attached.

## Data Availability

All data used in the present study are publicly available for registered researchers.

https://hrsdata.isr.umich.edu/

https://share-project.org

https://www.elsa-project.ac.uk

https://www.rand.org/well-being/social-and-behavioral-policy/data/FLS/IFLS.html

https://charls.pku.edu.cn/en/

## Acknowledgement

I thank the participants in 31 countries around the world for providing information, time and in many visits blood biomarkers too, as well as the generous funding bodies over many decades. **HRS**: The HRS (Health and Retirement Study) is sponsored by the National Institute on Aging (grant number NIA U01AG009740) and is conducted by the University of Michigan. **ELSA**: The English Longitudinal Study of Ageing was developed by a team of researchers based at University College London, NatCen Social Research, the Institute for Fiscal Studies, the University of Manchester and the University of East Anglia. The data were collected by NatCen Social Research. The funding is currently provided by the National Institute on Aging in the US (grant numbers: 2RO1AG7644 and 2RO1AG017644-01A1), and a consortium of UK government departments coordinated by the National Institute for Health Research. **SHARE**: The SHARE data collection has been funded by the European Commission, DG RTD through FP5 (QLK6-CT-2001-00360), FP6 (SHARE-I3: RII-CT-2006-062193, COMPARE: CIT5-CT-2005-028857, SHARELIFE: CIT4-CT-2006-028812), FP7 (SHARE-PREP: GA N°211909, SHARE-LEAP: GA N°227822, SHARE M4: GA N°261982, DASISH: GA N°283646) and Horizon 2020 (SHARE-DEV3: GA N°676536, SHARE-COHESION: GA N°870628, SERISS: GA N°654221, SSHOC: GA N°823782, SHARE-COVID19: GA N°101015924) and by DG Employment, Social Affairs & Inclusion through VS 2015/0195, VS 2016/0135, VS 2018/0285, VS 2019/0332, and VS 2020/0313. Additional funding from the German Ministry of Education and Research, the Max Planck Society for the Advancement of Science, the U.S. National Institute on Aging (U01_AG09740-13S2, P01_AG005842, P01_AG08291, P30_AG12815, R21_AG025169, Y1-AG-4553-01, IAG_BSR06-11, OGHA_04-064, HHSN271201300071C, RAG052527A) and from various national funding sources is gratefully acknowledged (see www.share-project.org). CHARLS: the CHARLS research and field team coordinated by Peking University. IFLS: the RAND Corporation, University of Indonesia, University of Gadjah Mada, SurveyMETER Yogyakarta.

